# Similar effect of lidocaine and saline on ciliary beating of nasal epithelial cells in vitro

**DOI:** 10.1101/2024.03.25.24304837

**Authors:** Sibel T Savas, Stefan A Tschanz, Philipp Latzin, Carmen Casaulta, Loretta Müller

## Abstract

**Rationale:** Primary ciliary dyskinesia is a rare genetic disease affecting ciliary motility and causing respiratory symptoms. Diagnosis can be done by high-speed- videomicroscopy using nasal epithelial cells (NECs) obtained via brushings. This procedure can be painful, especially for children. The use of lidocaine is proposed to reduce this pain; however, it is not clear whether lidocaine changes ciliary beating frequency (CBF) or pattern (CBP) in the subsequent high-speed-videomicroscopy.

**Objective:** The aim of this study was to analyse the effect of lidocaine on the CBF and the CBP of differentiated, air-liquid-interface cultured NECs.

**Methods:** NECs from healthy volunteers were obtained via brushings and cultured at the air-liquid-interface. After differentiation, lidocaine or isotonic saline (IS, control) were added apically for 1 or 5 min each and CBF (in top view of whole inserts and side views of scratched cells) and CBP (only side view) were assessed and recorded up to 150 min. CBF was computed and CBP was analysed semiquantitatively.

**Results:** Lidocaine as well as IS increased the CBF in the top view approach significantly compared to baseline. However, we found no significant differences between lidocaine and IS (control) treatment. Additionally, no effect of lidocaine on CBF, CBP, amplitude, inter- and intracellular coordination or transport was seen in the side view approach.

## Conclusion

We conclude that the observed CBF increase is related to the addition of liquid on the mucus layer and not by the lidocaine itself. Therefore, it seems possible to use lidocaine for nasal analgesia without impact on subsequent analysis of the ciliary motility.

## 1. Introduction

Human airway epithelial cells are helpful in many diagnostic procedures as well as in research settings. They can be obtained via biopsies during bronchoscopies. However, this is quite an invasive method. Therefore, the less invasive nasal brushing has been established as a good alternative to obtain airway epithelial cells [1–3], e.g. for the diagnostics of primary ciliary dyskinesia (PCD) [4] or research about cystic fibrosis [5,6]. For PCD diagnostics, primary nasal epithelial cells (NECs) can either be analysed freshly or cultured at the air-liquid-interface (ALI), closely mimicking the conducting upper airways - with a pseudostratified epithelium containing mucus-producing and ciliated cells [7–12]. With the implementation of ALI cell cultures in the diagnostic workflow of PCD, the majority of the nasal brushings can be successfully cultivated, and thus re-brushings can be minimised to few cases with no successful cell cultivation [4,8]. For the diagnosis of PCD, the dynamics of the cilia of fresh brushings or scratched-off cell cultures are analysed with high-speed videomicroscopy (HSVM) regarding ciliary beating frequency (CBF) and pattern (CBP) [8,13]. HSVM is a recognised method for PCD diagnostics in Europe [4,8,13,14], however, it is still far from being standardised and several topics, such as ideal temperature or quantitative CBP evaluation, are still subject of current debate [15,16].

A nasal brushing is uncomfortable and can be painful, being of special consideration in children. One brushing, or even several re-brushings, can be perceived differently, ranging from indifference up to refusal to undergo the procedure due to painful experiences. Therefore, the use of locally applied anaesthesia has been discussed to mitigate some of the pain and could reduce the fear of the brushing. This procedure is already used in clinical practice before bronchoscopy, nasal septal surgery, manipulation of nasal fractures, endoscopic sinus surgery or nasogastric tube insertion, albeit here with questionable clinical relevance, especially in the paediatric population [17–23].

However, concerning PCD diagnostics, it is not only important to avoid pain during the brushings, it also has to be ensured that the use of local anaesthetics does not impact the quality of the diagnostic analysis via affecting the CBF or the CBP assessed in HSVM. Older publications suggest that lidocaine in higher concentrations affects the CBF and the ciliary beat harmony [24–28] and would make the use of lidocaine in the context of PCD diagnostics impossible. However, another publication showed no difference in CBF after in vivo application [25]. Therefore, the aim of this study was to investigate whether lidocaine at a concentration used for local anaesthesia changes CBF or CBP of nasal cilia.

## 2. Methods

### 2.1. Experimental setup

NECs obtained via nasal brushing from healthy volunteers were cultured at the ALI until fully differentiated. Lidocaine or isotonic saline (IS) as a control were applied apically and washed off after either 1 or 5 min. In the following, we performed two different types of analysis: (1) **Top view analysis** in the intact cell culture on the transwell (as previously described [6], N=8). This methods allows to most closely mimic what happens in vivo after nasal application of lidocaine since the cells stay in ALI conditions. The only readout of this approach is the CBF over time. (2) **Side view analysis** of scratched-off cells according to classical HSVM used for PCD diagnostics (for details see [8], N=5). This approach allows – since the cells can be observed from the side – to also assess the CBP, amplitude and coordination (in addition to CBF). However, the disadvantage of this approach is that the cells are submerged in media what does not represent physiological conditions. For both types of analyses the ciliary activity was followed over time.

### 2.2. Sampling of epithelial cells via nasal brushing

NECs were obtained from healthy adult volunteers via nasal brushings. After participants have blown their nose with a paper tissue, NECs were obtained by nasal brushings using one pre-wetted (0.9% NaCl) interdental brush (IDB-G50 3mm, Top Caredent, Zurich, Switzerland; elongated by attaching a 200μl pipette tip with parafilm) for each nostril, as previously described and evaluated [8,29]. Briefly, the brush is inserted into and removed out of the inferior turbinate of the nasal cavity a couple of times, while also slightly pressing against the lateral edges of the nasal cavity and rotating the brush. Brushes with obtained cells are immediately stored in a 15ml tube with media (RPMI, Sigma-Aldrich, cat no. R8758-500ML). The samples were cultured at the ALI, according to our previously published protocol [8] (for details see also below).

The study was approved by the Ethics Committees of the University Children Hospital, Inselspital Bern and of the Canton Bern, Switzerland (reference number 2018-02155). Written informed consent was obtained from all volunteers.

### 2.3. Cultivation of nasal epithelial cells at the air-liquid-interface and preparation for experiments

NECs were first placed in a submerged culture using the PneumaCult-Expansion-Plus medium (Stemcell Technologies, cat#05040), then they were cultured at the ALI using the PneumaCult-ALI medium (Stemcell Technologies, cat #05001) and 12-well plates (Falcon 12-well plate, REF353043) with Transwell permeable inserts (Corning Transwell polyester membrane inserts, pore size 0.4μm, diameter 12mm, CLS3460- 48EA, Sigma-Aldrich) (for details see [8]). After at least four weeks at the ALI and full differentiation, the cell cultures represent a respiratory epithelium with mucus-producing, ciliated and basal cells.

For the top view CBF evaluation in the intact cell cultures, one day before the experiment, the NECs were washed for 15 min with 37°C phosphate-buffered saline (PBS), containing Mg^2+^ and Ca^2+^ (Dulbecco’s PBS with MgCl_2_ and CaCl_2_ (PBS++), Sigma D8537-500ml), to remove excess mucus. Then, the transwells were placed in a new plate and fresh medium was added. The surrounding empty wells were filled with 1 ml PBS each, to support equilibrium in humidity and temperature during experiments.

### 2.4. Exposure to Lidocaine

We exposed the NECs either for 1 or 5 min to lidocaine to mimic the application on the nasal epithelium either by using a nasal spray prior to the brushing (1 min treatment) or via the placement of a cotton pad into the nasal cavity for few minutes (5 min treatment). 200 μl Lidocaine (1% Lidocaine HCl, 10mg/ml, Bichsel, art.-no: 2057642) or 200 μl isotonic saline (80 mg NaCl in 10 ml water (Aqua ad injectabilita, Bichsel, FE1010221), ion concentrations analogous to Lidocaine solution) were warmed to 37°C and added to the fully mature and differentiated cell cultures. After 1 or 5 min, the liquid was removed, the cultures were washed with PBS (also warmed to 37°C) and ciliary beating was recorded.

### 2.5. Recording and Analysis

Ciliary motility was assessed at 37°C using an inverted Olympus IX73 light microscope, equipped with a heating plate or the cellVivo incubation system. Recordings were performed with a high-speed C-MOS camera (FLIR 3.2 MP Mono Grasshopper3 USB 3.0 Camera, Sony IMX252 chip) for 2 s at 300 frames per second, with a resolution of 480 x 640 pixels and saved as 8-bit grey level tagged image file format (TIF) image series.

We performed two different types of analysis: (1) **Top view analysis** in the intact cell culture on the transwell (as previously described [6]): Whole transwells of cell cultures of eight healthy volunteers and a 10x objective in phase-contrast mode were used. The videos of each sample were recorded in a systematic manner at the same three spots of the insert and the whole field of view was used for CBF calculations. The temperature was kept to 37°C and the cell cultures were placed back in the incubator between recordings. (2) **Side view analysis** (only for the 5 min treatment) of scratched-off cells according to classical HSVM used for PCD diagnostics (for details see [8]): Briefly, cell agglomerates from the treated cell cultures of five healthy volunteers were scratched off from the transwells and transferred together with culture medium onto a closed chamber built by a microscopy glass slide, a 200µm thick rubber distance ring (Grace Bio-Labs CoverWell™ imaging chambers with removed cover slip; cat no. 645401) and a glass cover slip (22×22 mm, DURAN GROUP, Wertheim, Germany, cat.no. 23550327) on top. This setup allowed to record the cells from the side for the analysis of the CBP. Videos of the cells were captured with a 40x objective in bright field mode. Between recordings, the slides were left at room temperature (23°C) to mimic conditions of the PCD diagnostic procedure (at that time).

For both types of analyses the following timepoints were used: baseline (before the application of any liquids), 1 min (only for the 1 min treatment), 5 min, 10 min, 15 min, 30 min, 60 min, 90 min, 120 min and 150 min. CBF (of top and side view recordings) was computated using a Fourier-Transformation-based algorithm of the Cilialyzer, a Python-based program [30]. CBF values are presented as the mean of all recorded videos. All assessments of CBF were done by the same observer. When analysing CBP (of the side view recordings), each video recorded was described and analysed by a scoring system developed by S. Tschanz (for details see [8]). CBP is semiquantitatively scored with 0-4, with four referencing a physiological, complete pattern and zero referencing no pattern at all. Amplitude and coordination were also scored semiquantitatively. For those parameters the mean of all 10 videos was calculated for each timepoint of each participant and the mean of all participants for each timepoint is presented. Transport of cellular debris was scored as observed (1) or not (0). The % of transport (as presented in Table 1) is the mean of those scores and represents the ratio of videos with visible transport. Again, all assessments were done by the same observer.

**Table 1.** Assessment of ciliary beating in classical HSVM using side views of scratched-off cells (side view approach).

| **control** | **5 min** | **10 min** | **15 min** | **30 min** | **60 min** | **90 min** | **120 min** | **150 min** |
| --- | --- | --- | --- | --- | --- | --- | --- | --- |
| CBP | 3.93<br>(3.7-4.0) | 3.63<br>(3.5-4.0) | 3.53<br>(2.0-4.0) | 3.70<br>(3.3-4.0) | 3.60<br>(2.7-4.0) | 3.87<br>(3.0-4.0) | 3.29<br>(2.7-4.0) | 3.60<br>(3.0-4.0) |
| amplitude | 3.83<br>(3.5-4.0) | 3.77<br>(3.7-4.0) | 3.53<br>(2.0-4.0) | 3.9<br>(3.7-4.0) | 3.80<br>(3.7-4.0) | 3.73<br>(3.3-4.0) | 3.92<br>(3.7-4.0) | 3.73<br>(3.3-4.0) |
| Intracellular coordination | 1.93<br>(1.7-2.0) | 2.00<br>(2.0-2.0) | 1.67<br>(1.0-2.0) | 1.93<br>(1.7-2.0) | 1.93<br>(1.7-2.0) | 2.00<br>(2.0-2.0) | 1.83<br>(1.0-1.7) | 1.93<br>(1.7-2.0) |
| Intercellular coordination | 1.20<br>(0.0-2.0) | 1.50<br>(0.3-2.0) | 1.27<br>(0.0-2.0) | 1.40<br>(1.0-2.0) | 1.60<br>(1.3-2.0) | 1.77<br>(1.3-2.0) | 1.50<br>(1.0-1.7) | 1.50<br>(1.0-1.7) |
| Transport (%) | 53<br>(67-100) | 63<br>(0-100) | 67<br>(33-100) | 87<br>(33-100) | 67<br>(0-100) | 67<br>(0-100) | 75<br>(33-100) | 80<br>(67-100) |
| CBF | 5.58±0.72<br>(4.7-6.3) | 5.72±0.92<br>(4.5-6.7) | 5.75±1.0<br>(4.6-7.1) | 5.41±0.88<br>(4.7-6.4) | 4.84±0.66<br>(4.2-5.9) | 4.96±0.8<br>(3.7-5.8) | 4.63±0.79<br>(3.8-5.9) | 5.15±1.54<br>(3.3-7.4) |
| **lidocaine** | **5 min** | **10 min** | **15 min** | **30 min** | **60 min** | **90 min** | **120 min** | **150 min** |
| CBP | 3.67<br>(3.0-4.0) | 3.88<br>(3.5-4.0) | 3.73<br>(3.0-4.0) | 3.33<br>(3.0-3.7) | 3.70<br>(3.3-4.0) | 3.58<br>(3.0-4.0) | 3.67<br>(3.3-4.0) | 3.33<br>(2.7-4.0) |
| amplitude | 3.92<br>(3.7-4.0) | 3.75<br>(3.3-4.0) | 3.80<br>(3.7-4.0) | 3.87<br>(3.3-4.0) | 3.57<br>(2.7-4.0) | 3.75<br>(3.3-4.0) | 3.83<br>(3.5-4.0) | 3.60<br>(3.0-4.0) |
| intracellular coordination | 2.00<br>(2.0-2.0) | 2.00<br>(2.0-2.0) | 1.93<br>(1.7-2.0) | 1.80<br>(1.0-2.0) | 2.00<br>(2.0-2.0) | 1.92<br>(1.7-2.0) | 1.93<br>(1.7-2.0) | 2.00<br>(2.0-2.0) |
| intercellular coordination | 1.71<br>(1.5-2.0) | 1.83<br>(1.7-2.0) | 1.53<br>(1.0-2.0) | 1.30<br>(0.5-1.7) | 1.63<br>(1.0-2.0) | 1.33<br>(0.7-2.0) | 1.40<br>(1.3-1.7) | 1.47<br>(0.3-2.0) |
| Transport (%) | 75<br>(0-100) | 58<br>(0-100) | 87<br>(33-100) | 77<br>(33-100) | 80<br>(33-100) | 63<br>(0-100) | 80<br>(33-100) | 87<br>(67-100) |
| CBF | 6.66±0.39<br>(6.4-7.1) | 6.23±0.84<br>(5.3-6.7) | 6.33±1.38<br>(4.7-8.0) | 5.74±0.79<br>(4.7-6.6) | 4.48±0.64<br>(3.8-4.9) | 5.12±0.58<br>(4.5-5.7) | 5.23±0.61<br>(4.9-6.0) | 4.75±1.11<br>(3.7-6.3) |

Data are presented as mean (range) for CBP, amplitude and coordination, as mean % (range) of videos with particle transport for transport and as mean ± standard deviation (range) for CBF.

### 2.6. Statistical Analysis

Data collection and statistics were done via Excel and GraphPad Prism Version 9.0.2 (GraphPad Software, San Diego, California, USA). Wilcoxon matched-pairs signed rank tests were performed (with absolute values only) to compare (1) all after-treatment timepoints to the corresponding baseline and (2) the two treatments at different corresponding timepoints. A p-value <0.05 was accepted as a statistically significant difference.

## 3. Results

### 3.1. Study Population

In total, eight healthy volunteers (five female, three male) were recruited. All participants stated to be non-smokers and free of respiratory symptoms for at last four weeks before the brushing was done. The mean age was 31.4 years with a range of 20 to 51 years.

### 3.2. Increased CBF by lidocaine and control treatment in the top view approach

Lidocaine as well as isotonic saline (control) increased the CBF significantly to almost double CBF compared to the baseline after 30min and stayed as high until the end of the recordings after 150 min (Figure 1). However, there was no difference between lidocaine and control treatments, neither in absolute values (Figure 1A) nor when looking at values normalised to the baseline (Figure 1B). The 1 min control started out at the baseline with a CBF of 6.93 Hz (range 5.07 - 9.73 Hz), reached 10.66 Hz (6.8 - 14.33 Hz) after 15 min and finally almost doubled with 11.77 Hz (8.7 - 15.8 Hz) after 150 min. While the lidocaine 1 min started with 5.42 Hz (3.93 - 8.8 Hz) at the baseline, increased to 11.2 Hz (9.5 - 13.9 Hz) after 15 min and was more than double of the baseline after 150 min with 11.54 Hz (8.57 - 14.6 Hz). The control of 5 min had a baseline CBF of 7.19 Hz (4.2 - 11.37 Hz), increased to 9.76 Hz (6.55 - 14.171Hz) after 15 min and reached 10.85 Hz (8.53 - 14.5 Hz) after 150 min. The 5 min lidocaine intervention started off at a baseline of 7.1 Hz (5.47 - 10.6 Hz), increased to 10.37 Hz (8.2 - 13.23 Hz) after 15 min and slightly increased further to 11.1 Hz (7.0 - 15.5 Hz) after 150 min.

**Figure 1.**
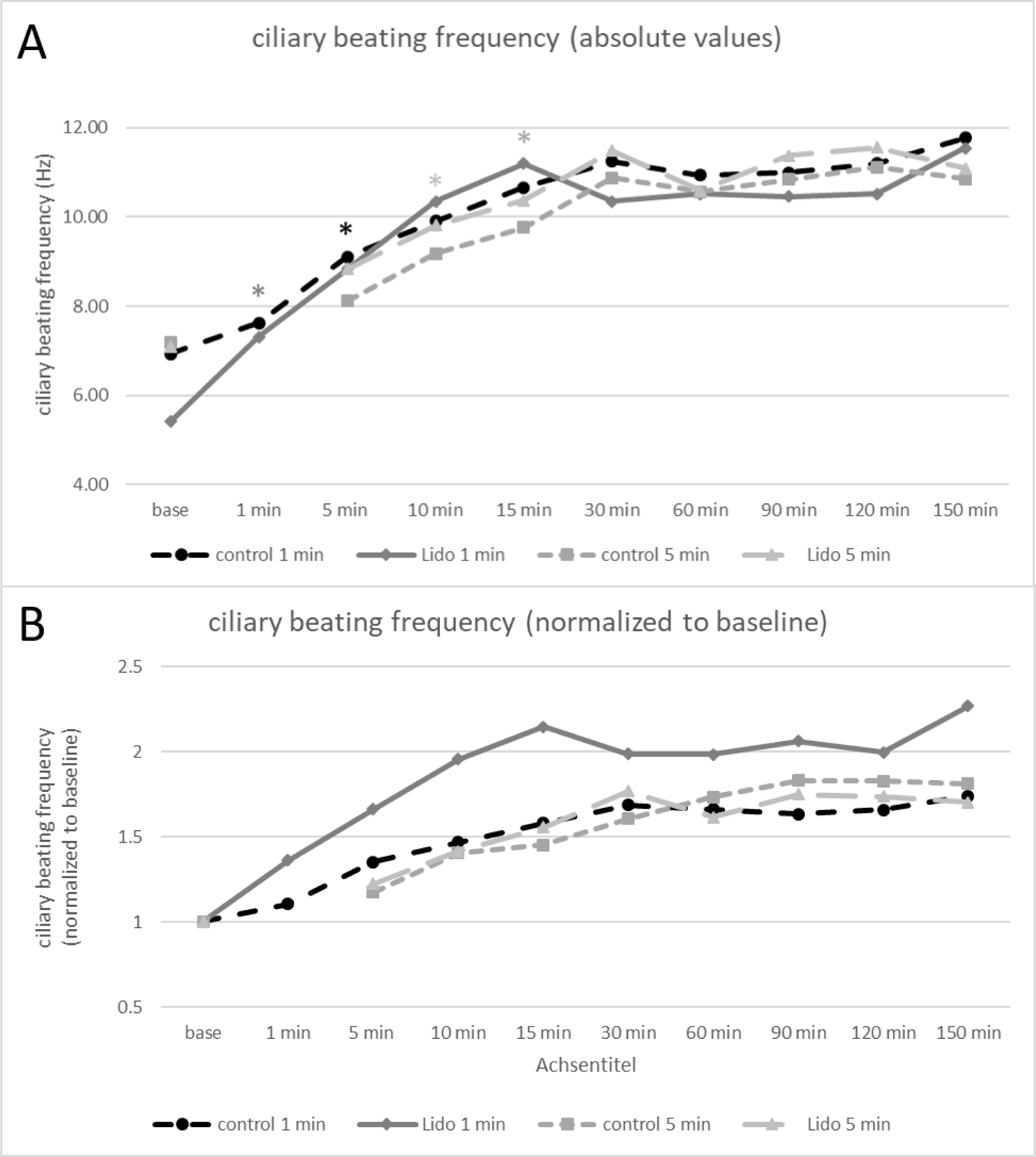
(A) Ciliary beating frequency over time after treatment of ALI cell cultures with lidocaine or saline (control). *p<0.05 with Wilcoxon matched-pairs signed rank test compared to baseline, once significant all later timepoints were also significantly different compared to baseline. (B) Ciliary beating frequency over time after treatment of ALI cell cultures with lidocaine or saline (control) for 1 or 5 min, normalized to baseline. Data are shown as means (of usually 10 videos) of all 8 donors.

When saline was applied for 1 min or 5min, the effect on CBF was statistically significant after 5 min or 15 min, respectively, and stayed until the end of the experiment (150 min). When lidocaine was applied for 1 min, the effect on CBF was statistically significant immediately after 1 min in comparison to baseline, when lidocaine was applied for 5 min, the effect was statistically significant in comparison to baseline after 10 min.

There were no statistically significant differences in CBF between the lidocaine and the control treatment with saline.

### 3.3. Similar effect of lidocaine and saline on ciliary beating pattern or frequency in the side view approach

There was no effect of lidocaine compared to the control treatment on the CBP, amplitude, intra- and intercellular coordination or transport in the side view approach after 5 min of liquid application (1 min application was not performed) (Table 1). <u>CBP</u> was 3.93 (range of 3.67 - 4.0) in the control group and 3.67 (3.0 - 4.0) in the lidocaine group 5 min after the treatment, and 3.6 (3.0 - 4.0) in the control group and 3.33 (2.67 - 4.0) in the lidocaine group after 150 min. The values were similar for the <u>amplitude</u> observed. <u>Intracellular coordination</u> remained very stable (5 min: 1.93 for control and 2.00 for lidocaine; 150 min: 1.93 for control, 2.0 for lidocaine), however <u>intercellular coordination</u> varied in both groups (5 min: 1.20 for control and 1.71 for lidocaine; 150 min: 1.50 for control, 1.47 for lidocaine). Transport was visible in most of the videos (5 min: 53% for control and 100% for lidocaine; 150 min: 80% for control, 87% for lidocaine). CBF in the side view approach was slightly decreasing over time, albeit not statistically significant (5.58 Hz at 5min to 5.15 Hz at 150min in the control group, 6.66 Hz to 4.75 Hz for the lidocaine group).

## 4. Discussion

We showed that treatment of ALI NEC cultures of healthy volunteers with lidocaine did not affect CBF differently than the control treatment. CBF increased similarly in whole inserts of cell cultures treated with either lidocaine or control saline to roughly twice the baseline values. The CBF, CBP, the amplitude and the coordination as well as particle transport did not differ in side view HSVM between cells treated with lidocaine or control saline.

The development of CBF values was different between the two approaches: CBF in the whole inserts with top view increased from roughly 7 to 11 Hz over the first 30min after the treatment. However, in the side view of the HSVM approach it decreased from roughly 7 to 5 Hz over 150min. There are several potential reasons for those differences. Frist of all, the top view recordings in the whole inserts were done at 37°C and the side view measurements at room temperature, approximately 23°C, with probably some drop of temperature since the cells and the media started at 37°C. As it is known that CBF highly depends on temperature, the temperature can explain the higher values of the top view approach and the decrease in the side view approach. Second, the side view samples are cell agglomerates in a closed imaging chamber submerged in media. Thus, those cells are surrounded with media, which represents an ending source of energy. In the side view approach, there is clearly less media per cell present compared to the whole insert top view approach. Therefore, in the side view approach exhaustion of media could be another reason for decreasing CBF over time. However, against this explanation speaks that the beating pattern was - even after 150min - still physiological. Strengths of our study include the closely mimicked in vivo conditions in vitro, such as the short application time, realistic concentrations as well as larger cell conglomerates. The ALI cultures nicely mimic the conducting airways [9–12] with the different epithelial cell types, including the ciliated cells of interest for this study. Additionally, obtaining CBF and CBP measurements from ALI cultures provides more reliable results than obtaining these values from freshly brushed cells since the quality of the sample is better and there are more cells and bigger cell groups to assess.

Limitations include that the lidocaine was not applied in vivo to the nose of healthy volunteers, but in vitro to cell cultures. We are aware that our study does not cover potential effects involving other respiratory cells than epithelial cells. Thus, future research should study the effects in vivo in a group of patients suspicious for PCD. Rutland et al. [25] have already studied the effect of lignocaine in vivo in heathy volunteers in 1982 and did not find any effects on CBF in brushed human nasal epithelium up to 24 hours after in vivo aerosolisation. Thus, our study supports these findings. Another limitation is the type of liquid application: a volume of 200 μl of liquid was added directly onto the cell layer and not nebulised, which would have more closely represented a nasal spray application. Even though the liquid was removed shortly afterwards (1 or 5 min later), this might have resulted in changes different than by nebulisation. Additionally, our cell cultures consist only of different types of epithelial cells (basal, mucus producing and ciliated cells) without other cell types that are usually present in the nose, such as immune cells.

In contrast to some of the rare published literature, we could not see the ciliodepressive or even ciliostatic effects that were described previously [24–28,31–33]. However, our study setting was also slightly different to the other studies. Mostow et al. incubated ferret trachea for 60 min with 1% and 2% lidocaine and observed complete ciliostasis, which was partially reversible at 1% after washing and removal of the trachea from the lidocaine incubation chamber. At lower concentrations (0.25 and 0.5%) they observed no effects [24]. Verra et al. applied lidocaine onto tracheal rings of guinea pigs and cattles where incubation with 1% lidocaine showed a marked decrease in CBF. However, the effect was shown to be completely reversible after rinsing [26]. Ingels et al. showed a decrease in CBF at 0.25% lidocaine application and irreversible ciliostasis at 2% in human nasal mucosa biopsy specimen. However, they used a superfusion chamber keeping the biopsies in submerged conditions and not at the ALI. In comparison to our setting this may mask any hydration effect of the mucus layer we have observed. The duration of the treatment was 6 min, thus quite similar to our study (1 or 5 min). Additionally, this publication did not report on the temperature, which can have a significant effect on ciliary motility [27]. Rutland, Griffin and Cole also showed a decreasing effect on CBF after adding lidocaine (lignocaine) at concentrations >0.25% to brushed human NECs ex vivo, progressing to ciliostasis when concentrations over 2% were applied. The samples were stored in media containing lignocaine for 30 min, not reflecting ALI conditions of the mucosa. They found a cilio- inhibition ex vivo, but only at concentrations far above from clinical practice and what we used in our study, however, as mentioned above, they did not find any effects on CBF up to 24 hours in brushed human nasal epithelium after in vivo aerosolisation [25]. We found a similar effect of lidocaine and control saline on the CBF and therefore conclude that the increased CBF most probably relates to the addition of fluid onto the periciliary layer and a hydration of the mucus layer [35]. The strong effect of mucus hydration due to adding liquid could maybe mask an inhibitory effect induced by lidocaine. However, in our study we wanted to test whether the application of lidocaine in the context of a nasal brushing for PCD diagnostics would affect the downstream analysis of CBF and CBP, and thus we can conclude here that our findings do not speak against the application of lidocaine to reduce pain of nasal brushings performed for the analysis of the ciliary motility.

We conclude that application of lidocaine in vitro does not affect the assessment of the ciliary motility via HSVM and that this may also be true for in vivo use in the context of nasal brushings as done for PCD diagnostics. This would be especially relevant for children that according to the ERS algorithm would have to be brushed three times if cell culture is not available at the diagnostic centre they’re referred to.

## Data Availability

All data produced in the present study are available upon reasonable request to the authors.

## Acknowledgments

We thank all volunteers for donating nasal cells and Andrea Stokes (Division of Paediatric Respiratory Medicine and Allergology, Department of Paediatrics, Inselspital; Lung Precision Medicine (LPM), Department of BioMedical Research (DBMR), University of Bern, Switzerland) for her excellent lab work.

## Author contribution

STS helped with study design, performed all experiments, analysed the data and wrote the manuscript. SAT helped with study design and data analysis. PL helped with study design and data interpretation. CC came up with the original project idea, helped with study design and data interpretation. LM led the project, was responsible for the study design, helped with data analysis, data interpretation and wrote the manuscript. All authors read and approved the final version of this manuscript.

